# Usefulness of pystan and numpyro in Bayesian item response theory

**DOI:** 10.1101/2023.03.29.23287903

**Authors:** Mizuho Nishio, Eiji Ota, Hidetoshi Matsuo, Takaaki Matsunaga, Aki Miyazaki, Takamichi Murakami

## Abstract

**Purpose:** The purpose of this study is to compare two libraries dedicated to Markov chain Monte Carlo method: pystan and numpyro.

**Materials and methods:** Bayesian item response theory (IRT), 1PL-IRT and 2PL-IRT, were implemented with pystan and numpyro. Then, the Bayesian 1PL-IRT and 2PL-IRT were applied to two types of medical data obtained from a published paper. The same prior distributions of latent parameters were used in both pystan and numpyro. Estimation results of latent parameters of 1PL-IRT and 2PL-IRT were compared between pystan and numpyro. Additionally, the computational cost of Markov chain Monte Carlo method was compared between the two libraries. To evaluate the computational cost of IRT models, simulation data were generated from the medical data and numpyro.

**Results:** For all the combinations of IRT types (1PL-IRT or 2PL-IRT) and medical data types, the mean and standard deviation of the estimated latent parameters were in good agreement between pystan and numpyro. In most cases, the sampling time using Markov chain Monte Carlo method was shorter in numpyro than that in pystan. When the large-sized simulation data were used, numpyro with a graphics processing unit was useful for reducing the sampling time.

**Conclusion:** Numpyro and pystan were useful for applying the Bayesian 1PL-IRT and 2PL-IRT.

## 1 Introduction

Item response theory (IRT) is a statistical framework used for analyzing test results and evaluating test items and test takers quantitatively. While IRT is commonly used in educational and psychological research (1–3), there are several applications of IRT to medical research. For example, Choi et al. used IRT for constructing the computer adaptive testing system of short-form patient-reported outcome measures with the data from the Patient-Reported Outcomes Measurement Information System project (4). Gershon et al. used IRT to build the quality of life item banks for adults with neurological disorders (5).

Generally, IRT is applied to the results of binary responses to the test items (e.g., correct and incorrect answers). In medical diagnosis, the results of various diagnostic procedures are frequently defined as binary responses. Therefore, it is possible to apply IRT to the data of medical diagnosis. To apply IRT to the data of medical diagnosis, the following correspondence is assumed: (i) the patient as the test item, (ii) the doctor as the test taker, and (iii) the results of the binary responses obtained through medical diagnosis as test results. For example, Nishio et al. used IRT for analyzing the results of medical diagnoses by radiologists (6).

The Bayesian IRT can be implemented using probabilistic programming languages or dedicated libraries (e.g., JAGS, Stan, pystan, and numpyro) (7–10). For example, previous studies used Stan for the implementation of the Bayesian IRT, graded response model, and nominal response model (6,11,12). The recent advances in hardware and software make it possible to use the Bayesian IRT efficiently. However, there is no study comparing the efficiency of the Bayesian IRT from the viewpoint of computational cost.

The purpose of the current study was to compare the results of the Bayesian IRT implemented with two dedicated libraries (pystan and numpyro). In the current study, the Bayesian 1PL-IRT and 2PL-IRT implemented with pystan and numpyro were applied to the two types of medical data obtained from the published paper (6). The estimation results of latent parameters and the computational cost were compared between pystan and numpyro. For reproducibility, our implementation of the Bayesian IRT in pystan and numpyro used in the current study is disclosed as open source through GitHub (https://github.com/jurader/irt_pystan_numpyro).

## 2 Materials and methods

Because this study used the medical data obtained from the published paper, institutional review board approval or informed consent of patients was not necessary.

### 2.1 Medical Data

The two types of medical data (BONE and BRAIN data) were obtained from the published paper (6). Table 1 shows the characteristics of the two types of medical data. The BONE data include binary responses from 60 patients (test items) and 7 radiologists (test takers), and the BRAIN data include those from 42 patients and 14 radiologists. The total numbers of the binary responses were 420 and 588 in the BONE and BRAIN data, respectively.

**Table 1.** Characteristics of two types of medical data.

| Name of data | Number of test takers | Number of test items | Total number of<br>binary responses |
| --- | --- | --- | --- |
| BONE data | 7 | 60 | 420 |
| BRAIN data | 14 | 42 | 588 |

### 2.2 1PL-IRT

IRT is a statistical model for analyzing the results of binary responses. While there are several types of IRT models (13), 1PL-IRT and 2PL-IRT were used. In the current study, latent parameters of IRT are estimated based on the results of medical diagnoses by test takers.

In 1PL-IRT, one latent parameter (β_*i*_) is used to represent the difficulty of test item *i*, and another latent parameter (θ_*j*_) is used to represent the ability of test taker *j*. The following equations represent 1PL-IRT.

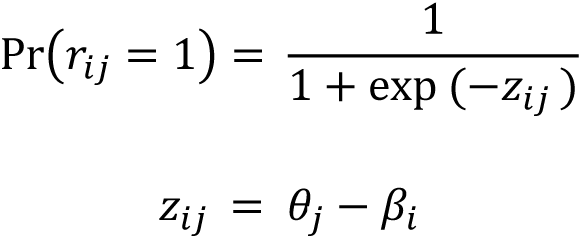

Here,

- Pr(*r*_*ij*_ = 1) represents the probability that the response of test taker *j* to test item *i* is correct,
- β_*i*_ is the difficulty parameter of test item *i*,
- θ_*j*_ is the ability parameter of test taker *j*.

### 2.3 2PL-IRT

In 2PL-IRT, two latent parameters (α_*i*_ and β_*i*_) are used to represent test item *i*. The following equations represent 2PL-IRT.

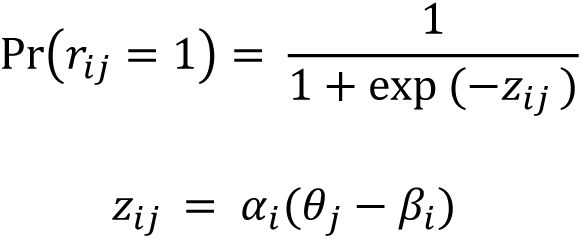

Here,

- α_*i*_ and β_*i*_ are the discrimination and difficulty parameters of test item *i*.

### 2.4 Experiments

We used Google Colaboratory to run the experiments. The following software packages were used on Google Colaboratory: pystan, version 3.3.0; jax, version 0.4.4; jaxlib, version 0.4.4+cuda11.cudnn82; numpyro, version 0.10.1. Two cores of Intel(R) Xeon(R) (2.20 GHz) and NVIDIA(R) Tesla T4(R) were used as CPU and a graphics processing unit (GPU), respectively.

#### 2.4.1 Experiments for agreement of latent parameters

The Bayesian 1PL-IRT and 2PL-IRT, implemented with pystan and numpyro, were applied to the BONE and BRAIN data. For 1PL-IRT, the following prior distributions were used:

- β_*i*_ ∼ *N*(0, 2),
- θ_*j*_ ∼ *N*(0, 2),

where *N* represents a normal distribution in which the first and second arguments are the average and variance of normal distribution, respectively. For 2PL-IRT, the following prior distribution was used in addition to those of 1PL-IRT:

- *log*(α_*i*_)∼ *N*(0.5, 1).

The same prior distributions of the latent parameters were used in both pystan and numpyro. The following parameters were used for sampling using Markov chain Monte Carlo method in both pytan and numpyro: num_chains=6, num_samples=8000, num_warmup=2000. For numpyro GPU version, chain_method=’parallel’ was used. After the sampling, the posterior distributions of the latent parameters were obtained for the Bayesian 1PL-IRT and 2PL-IRT. The posterior distributions of the latent parameters were then compared between pystan and numpyro.

#### 2.4.2 Experiments for computational cost

To evaluate the computation cost of 1PL-IRT and 2PL-IRT implemented with pystan and numpyro, simulation data were generated from the medical data (BONE and BRAIN data) and numpyro. The computer simulation was performed in the following steps: (i) estimating the posterior distributions of the latent parameters of 1PL-IRT and 2PL-IRT for the two types of medical data, and (ii) generating binary responses from the IRT equations and the estimated posterior distributions for the two types of medical data. The total number of binary responses in the simulation data were as follows: 420, 840, 2100, 4200, 8400, 21000, 42000, 84000, 210000, and 420000 for the BONE data; 588, 1176, 2940, 5880, 11760, 29400, 58800, 117600, 294000, and 588000 for the BRAIN data. To evaluate the computational cost in pystan and numpyro, the sampling time using Markov chain Monte Carlo method was measured. In numpyro, both CPU version and GPU version were used for the sampling. The following parameters were used for the sampling in both pytan and numpyro: num_chains=2, num_samples=3000, num_warmup=500. For numpyro GPU version, chain_method=’parallel’ was used. Due to the limitation of Google Colaboratory, it was not possible to evaluate the sampling time in several large-sized simulation data.

## 3 Results

In the current study, we focused on the ability parameters of test takers, and the estimation results of test items were omitted. Tables 2–5 present the estimation results of the ability parameters of test takers. In addition, Figures 1 and 2 show representative scatter plots of the estimation results between pytan and numpyro, which are obtained from values of Tables 2 and 5, respectively. Tables 2–5 show the mean, standard deviation, and credible interval (94% highest density interval) as the estimation results of the ability parameters of test takers. Based on Tables 2–5 and Figures 1 and 2, we found that there was good agreement between pystan and numpyro for 1PL-IRT and 2PL-IRT of the BONE and BRAIN data.

**Figure 1.**
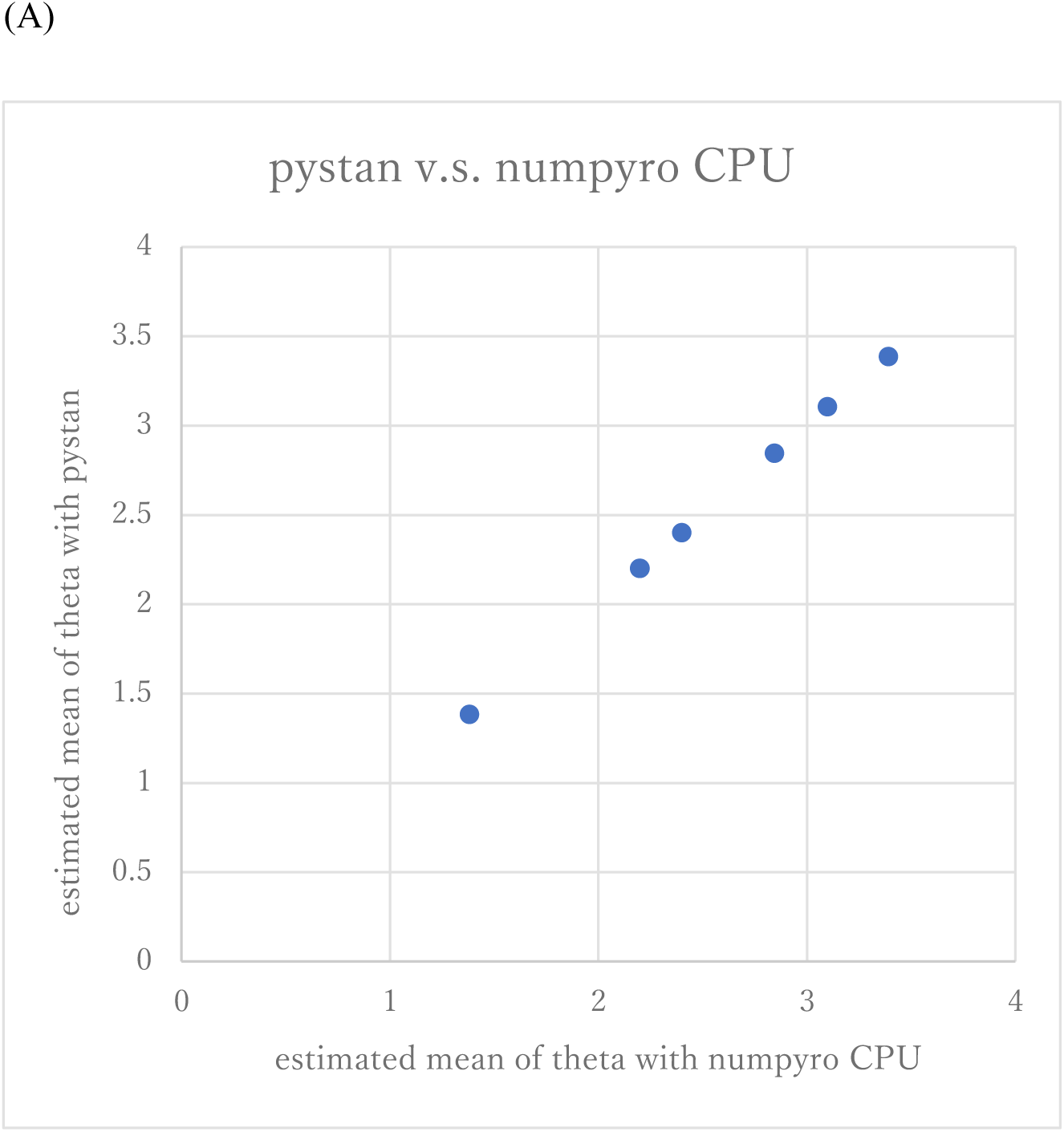
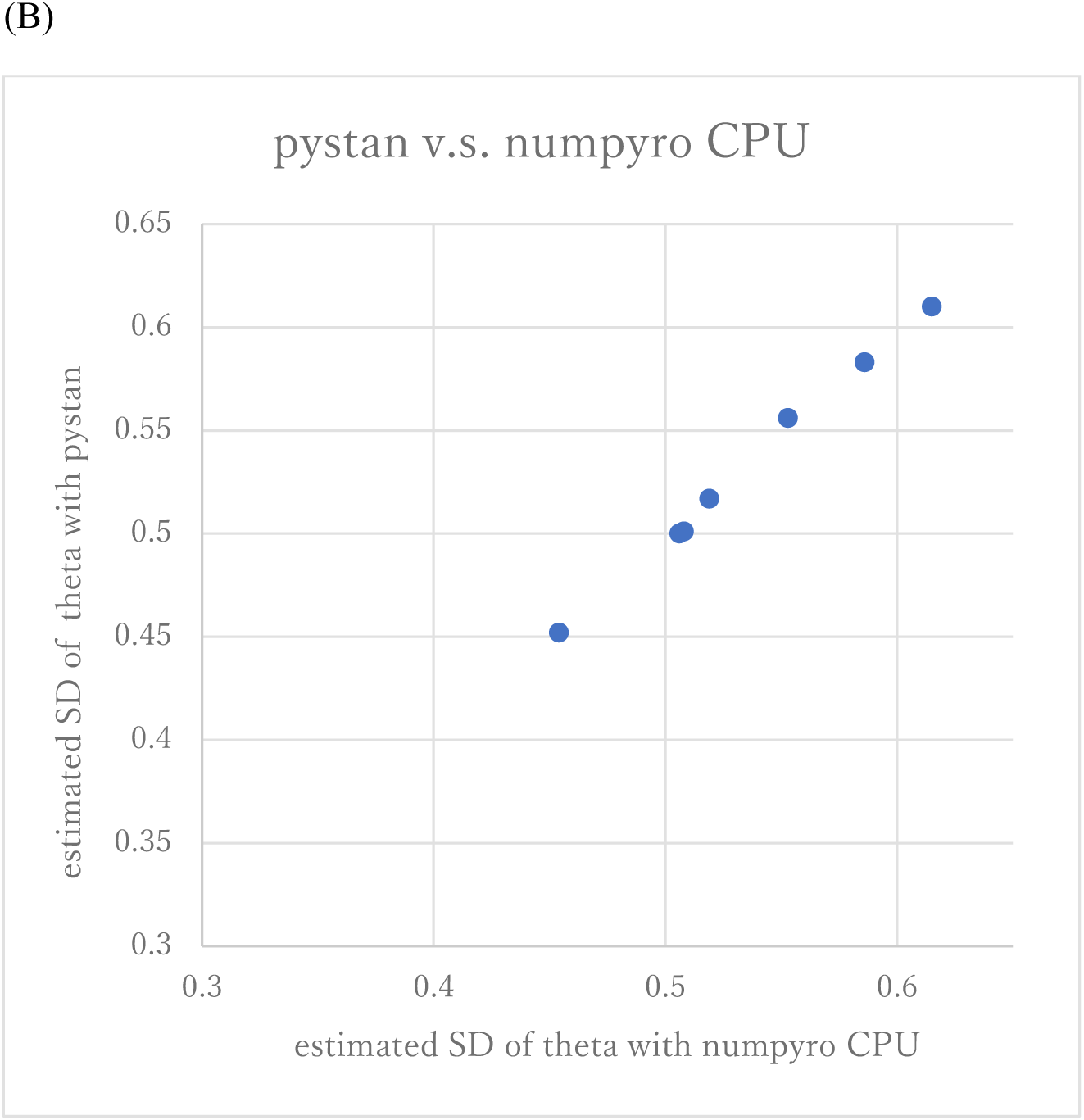
Representative scatter plots of estimated ability parameters of 1PL-IRT between numpyro and pystan for BONE data. (A) plot for mean of estimated ability parameters, (B) plot for SD of estimated ability parameters. Note: (A) and (B) are obtained from values of Table 2.

**Figure 2.**
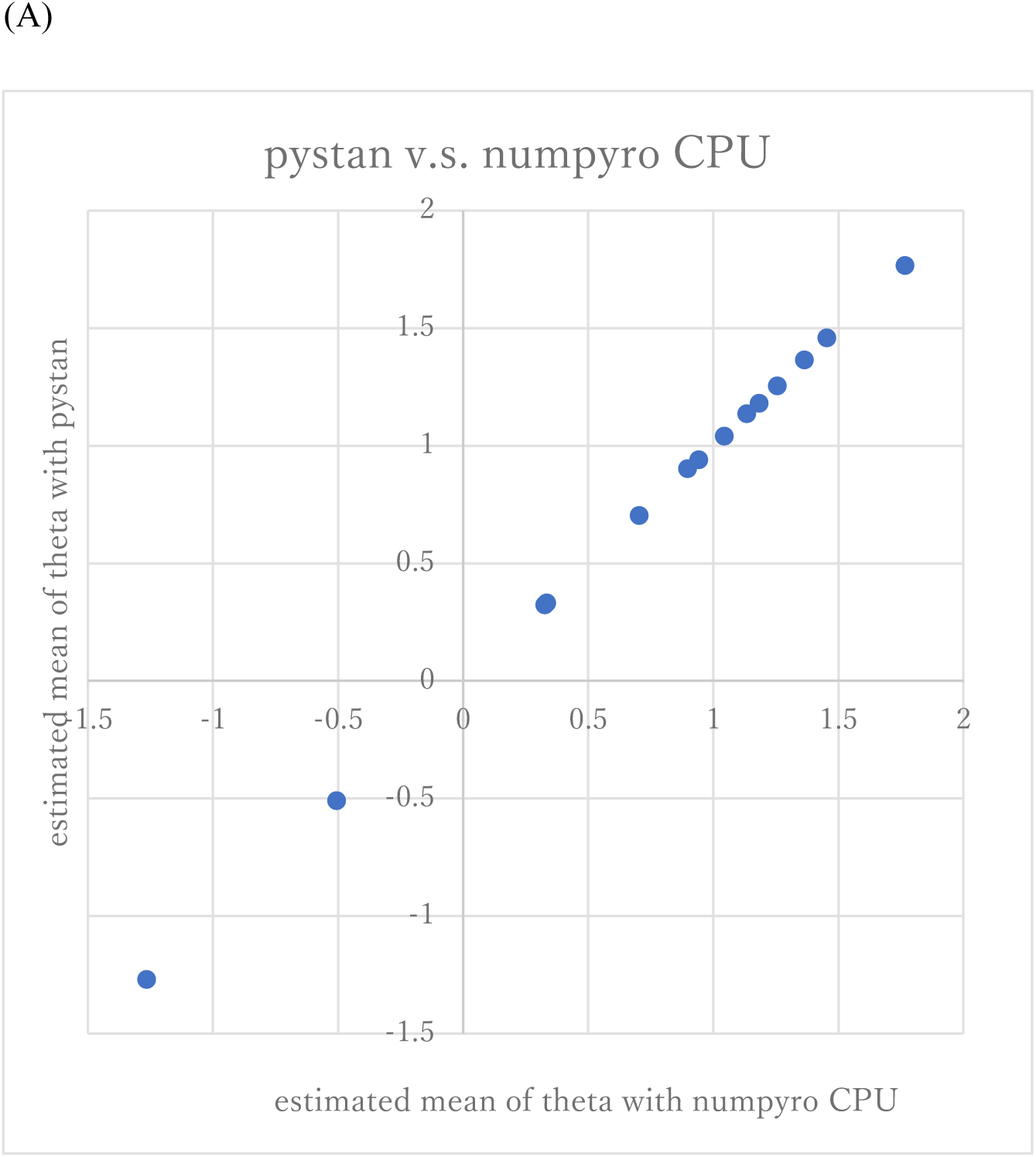
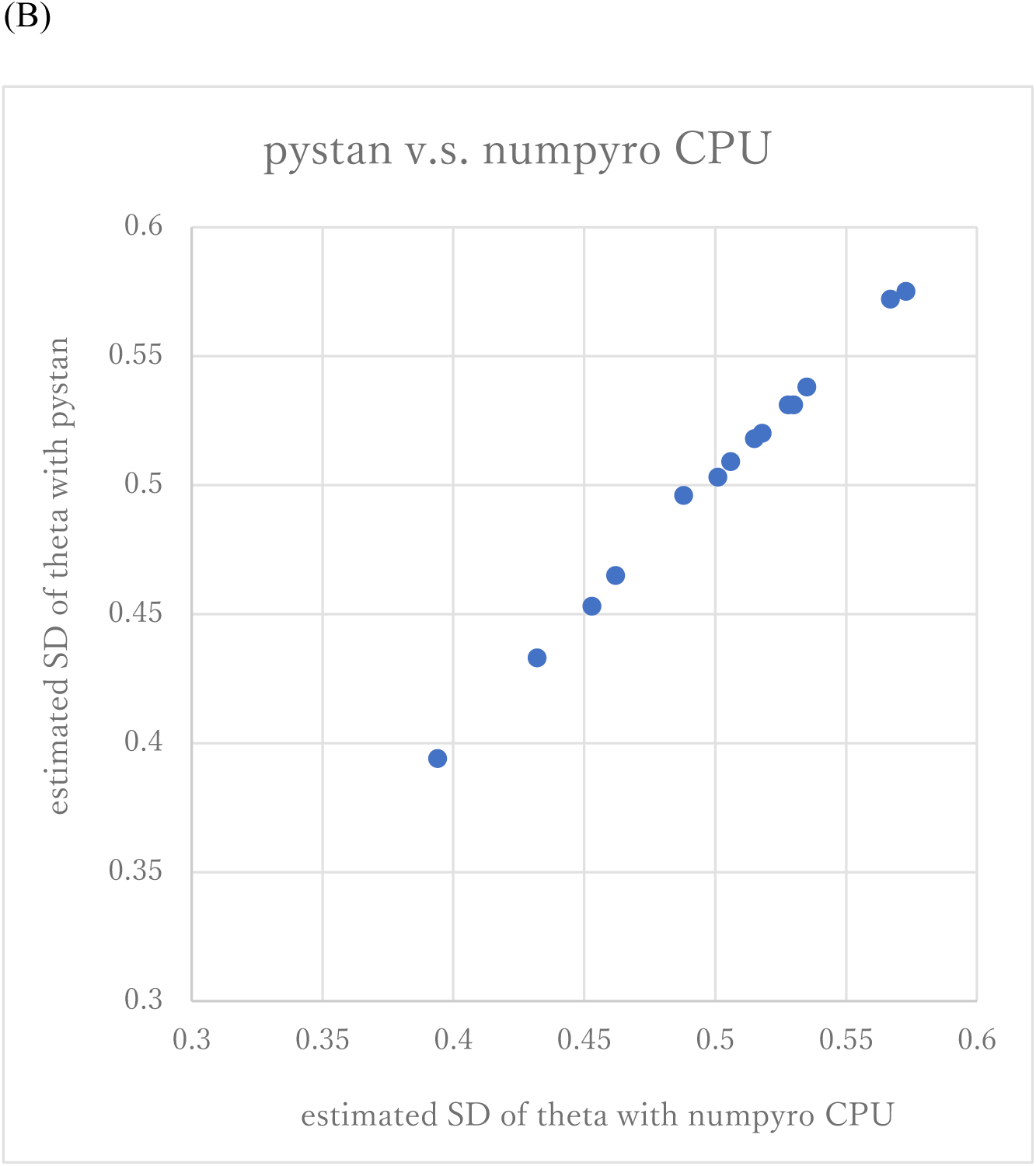
Representative scatter plots of estimated ability parameters of 2PL-IRT between numpyro and pystan for BONE data. (A) plot for mean of estimated ability parameters, (B) plot for SD of estimated ability parameters. Note: (A) and (B) are obtained from values of Table 5.

**Table 2.**
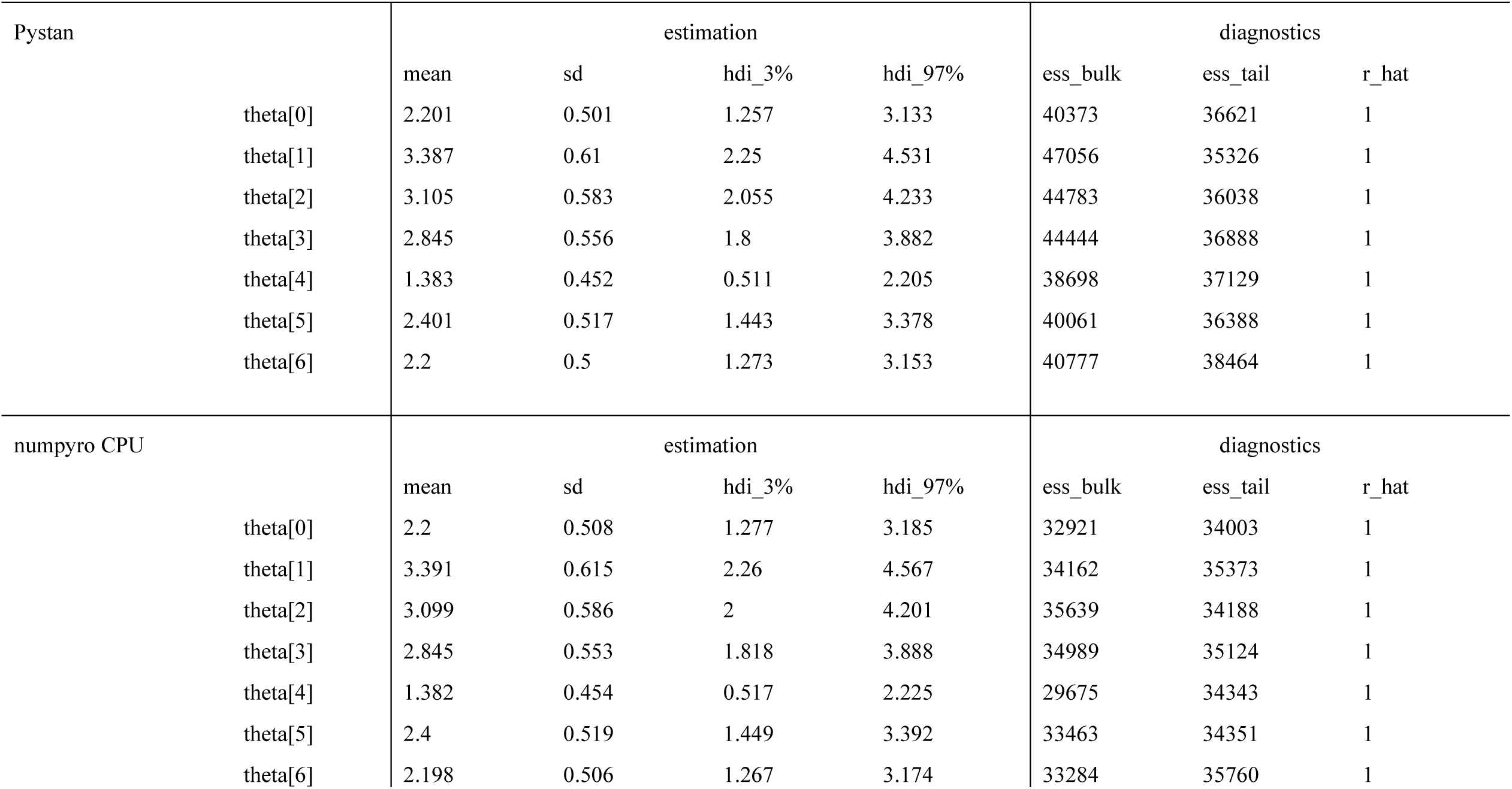
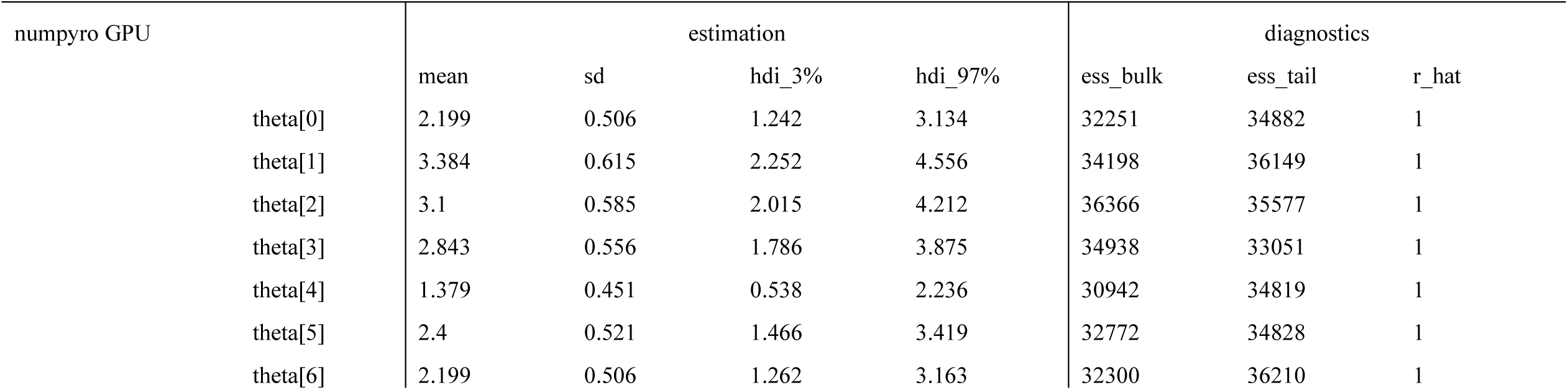
Estimation results of ability parameters of test takers in 1PL-IRT for BONE data.

**Table 3.** Estimation results of ability parameters of test takers in 1PL-IRT for BRAIN data.

| pystan | estimation |  |  |  | diagnostics |  |  |
| --- | --- | --- | --- | --- | --- | --- | --- |
|  | mean | sd | hdi_3% | hdi_97% | ess_bulk | ess_tail | r_hat |
| theta[0] | 2.062 | 0.49 | 1.128 | 2.976 | 14702 | 24025 | 1 |
| theta[1] | 3.086 | 0.605 | 1.966 | 4.231 | 17011 | 25730 | 1 |
| theta[2] | 2.677 | 0.539 | 1.664 | 3.687 | 16745 | 26368 | 1 |
| theta[3] | 2.585 | 0.571 | 1.534 | 3.658 | 16199 | 25432 | 1 |
| theta[4] | 1.252 | 0.412 | 0.482 | 2.029 | 14322 | 22764 | 1 |
| theta[5] | 2.096 | 0.536 | 1.102 | 3.111 | 13843 | 23047 | 1 |
| theta[6] | 2.169 | 0.511 | 1.206 | 3.125 | 15188 | 26449 | 1 |
| numpyro CPU | estimation |  |  |  | diagnostics |  |  |
|  | mean | sd | hdi_3% | hdi_97% | ess_bulk | ess_tail | r_hat |
| theta[0] | 2.057 | 0.486 | 1.159 | 2.991 | 15715 | 25398 | 1 |
| theta[1] | 3.086 | 0.607 | 1.943 | 4.196 | 17702 | 27769 | 1 |
| theta[2] | 2.676 | 0.546 | 1.679 | 3.723 | 16704 | 26124 | 1 |
| theta[3] | 2.581 | 0.576 | 1.515 | 3.674 | 15881 | 25012 | 1 |
| theta[4] | 1.248 | 0.409 | 0.494 | 2.037 | 15187 | 26085 | 1 |
| theta[5] | 2.095 | 0.533 | 1.106 | 3.094 | 15093 | 24705 | 1 |
| theta[6] | 2.167 | 0.511 | 1.194 | 3.104 | 15545 | 27676 | 1 |

| numpyro GPU | estimation |  |  |  | diagnostics |  |  |
| --- | --- | --- | --- | --- | --- | --- | --- |
|  | mean | sd | hdi_3% | hdi_97% | ess_bulk | ess_tail | r_hat |
| theta[0] | 2.063 | 0.487 | 1.159 | 2.988 | 14397 | 25263 | 1 |
| theta[1] | 3.09 | 0.601 | 1.975 | 4.226 | 16891 | 27636 | 1 |
| theta[2] | 2.68 | 0.546 | 1.658 | 3.701 | 15578 | 27217 | 1 |
| theta[3] | 2.585 | 0.576 | 1.525 | 3.682 | 14965 | 24243 | 1 |
| theta[4] | 1.251 | 0.411 | 0.503 | 2.046 | 14136 | 23883 | 1 |
| theta[5] | 2.096 | 0.538 | 1.105 | 3.112 | 13277 | 23510 | 1 |
| theta[6] | 2.171 | 0.513 | 1.237 | 3.162 | 14117 | 24946 | 1 |

**Table 4.**
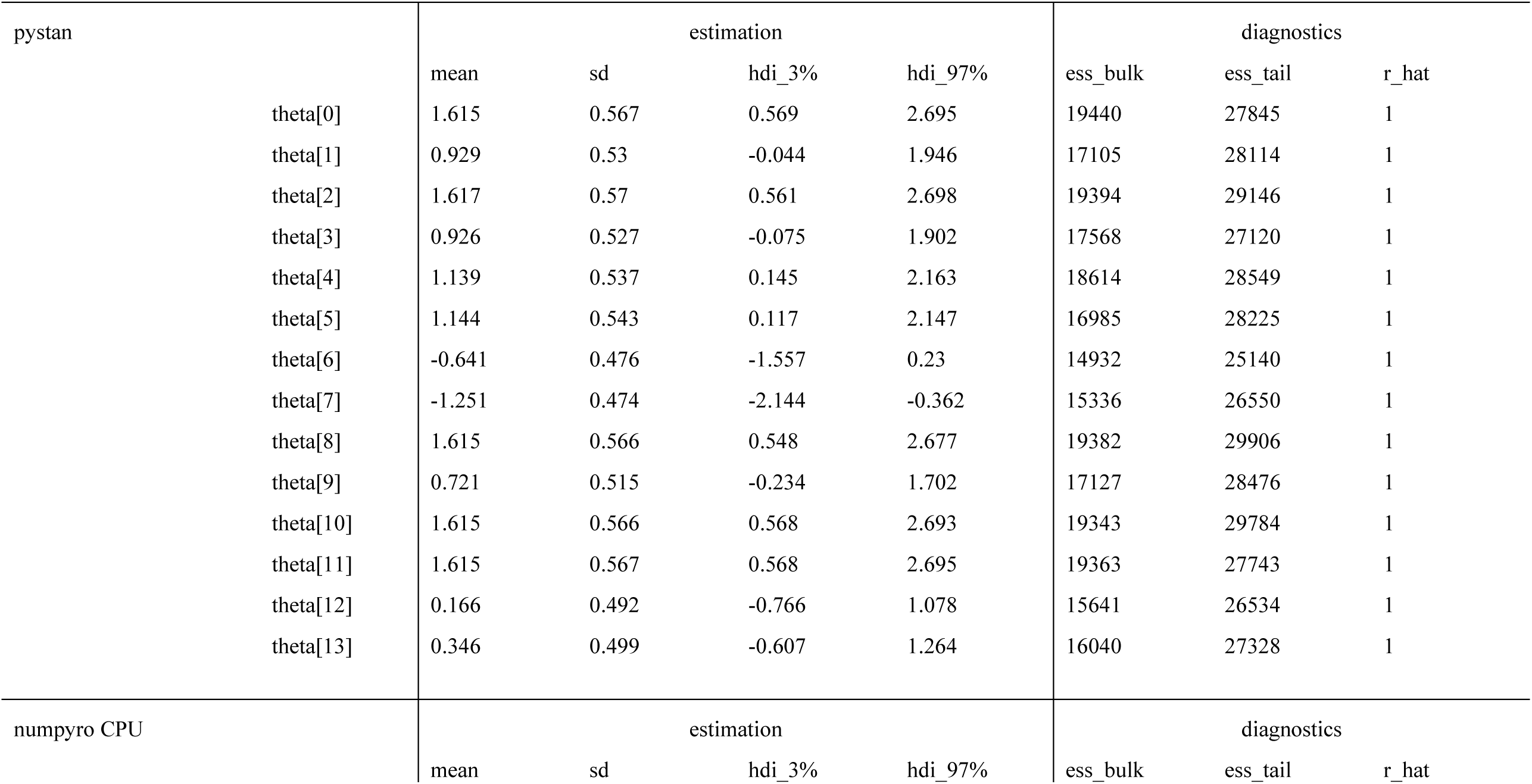
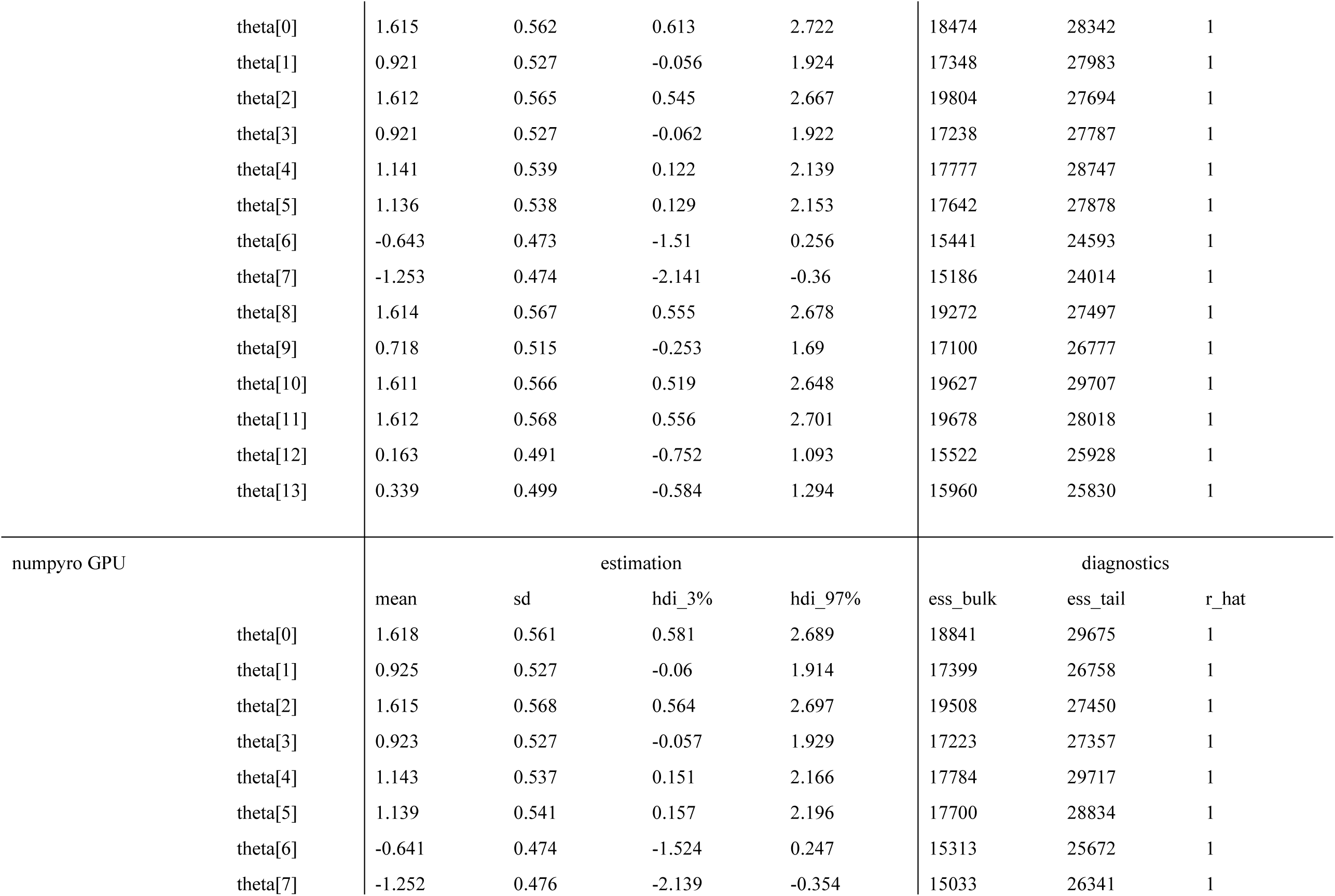
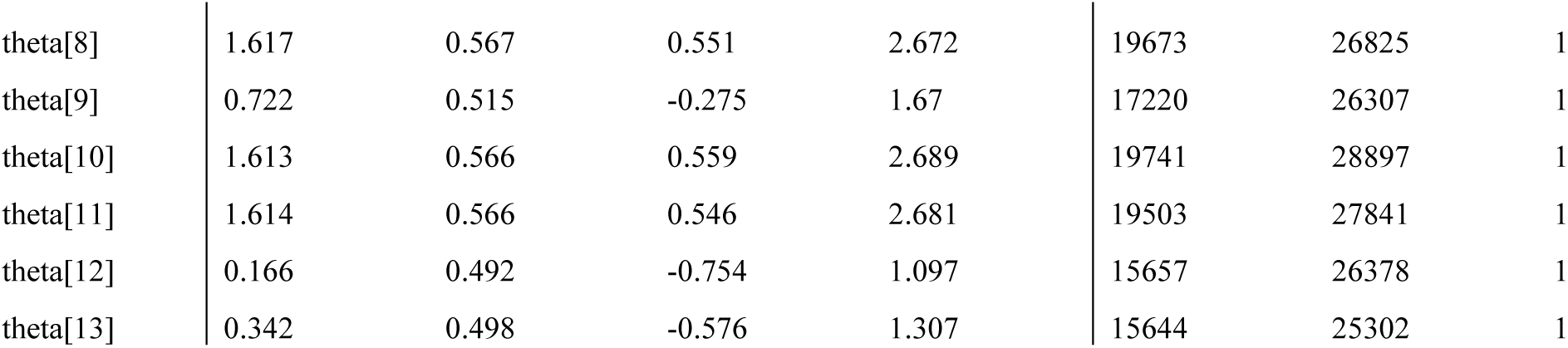
Estimation results of ability parameters of test takers in 2PL-IRT for BONE data.

**Table 5.**
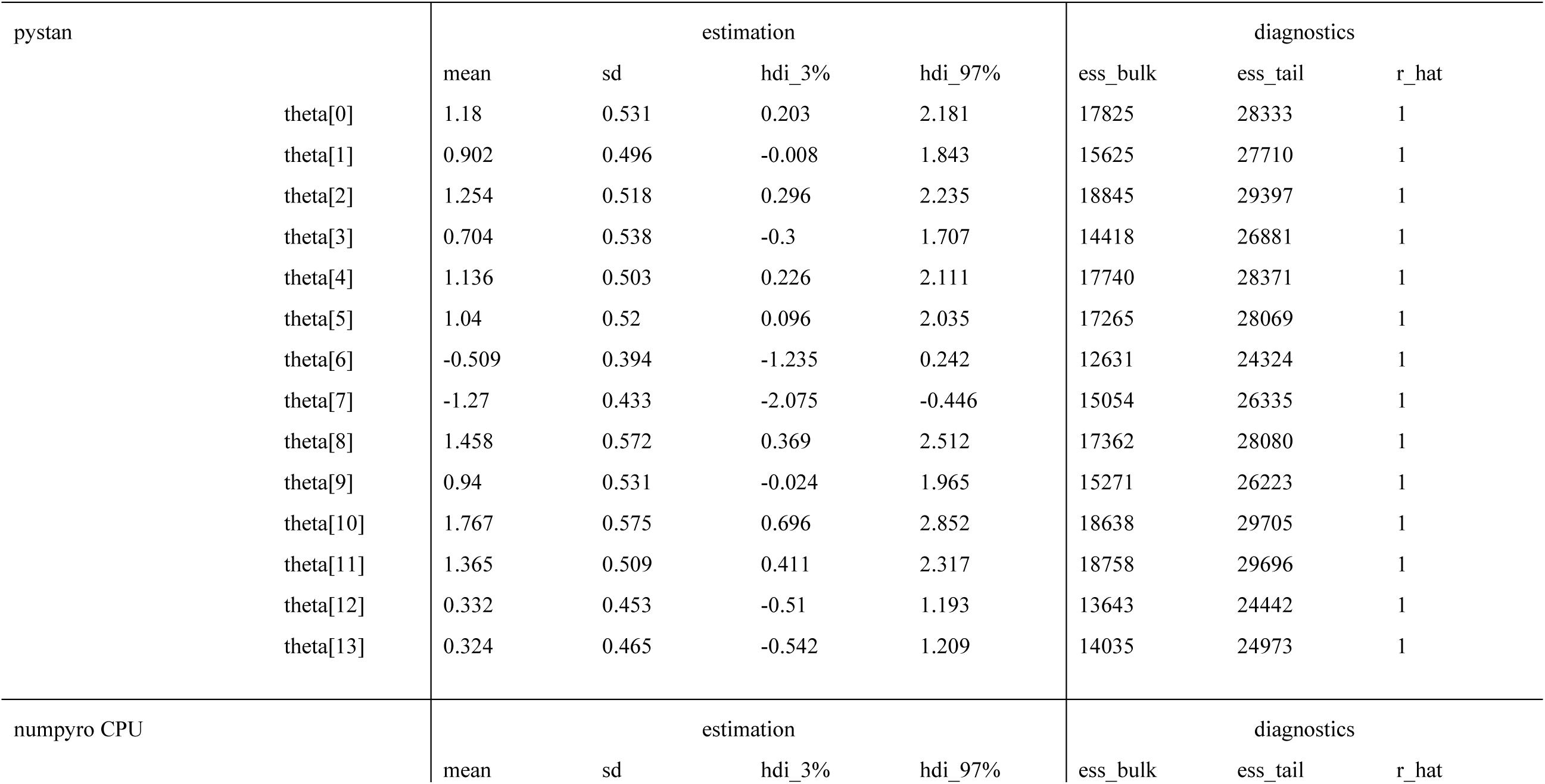
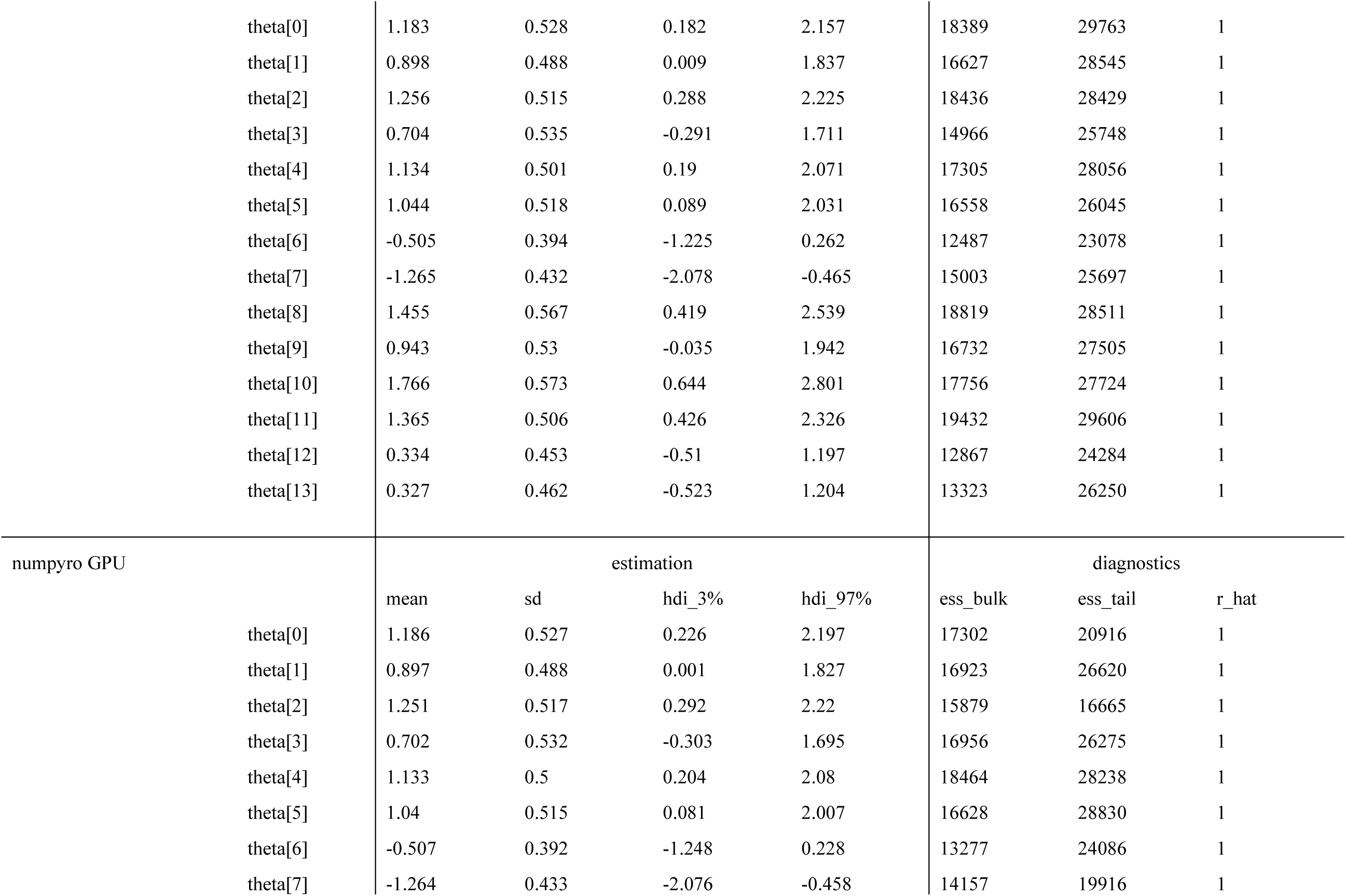
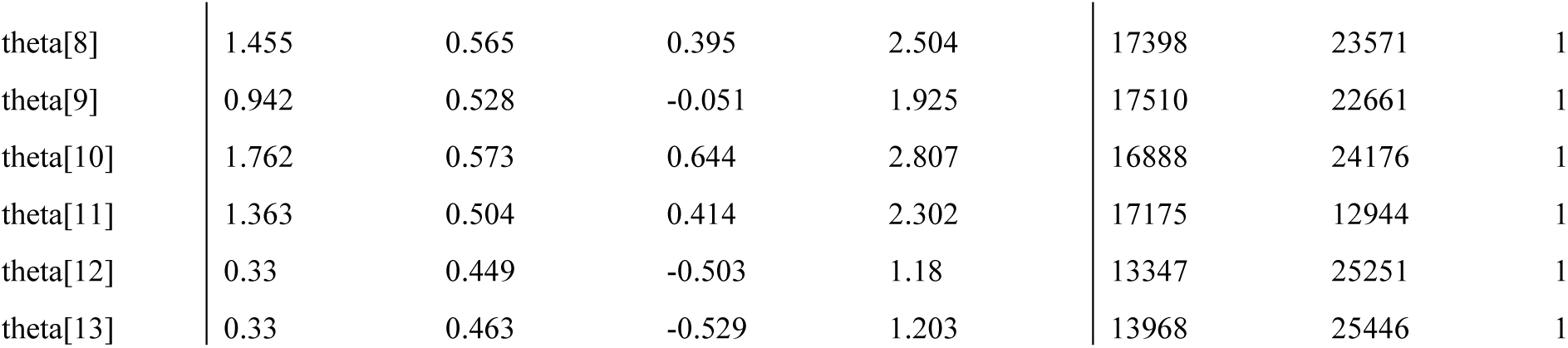
Estimation results of ability parameters of test takers in 2PL-IRT for BRAIN data.

From Tables 2–5, Lin’ s concordance correlation coefficients (CCC) of the estimated mean of the ability parameters were calculated between (a) pystan v.s. numpyro CPU version, (b) pystan v.s. numpyro GPU version, and (c) numpyro CPU version v.s. numpyro GPU version (14). The following criteria were used to evaluate CCC (15,16); low CCC values (< 0.900) were considered to represent poor agreement, whereas higher CCC values represented moderate (0.900–0.950), substantial (0.951–0.990), and almost perfect agreement (> 0.990). In the current study, the CCC values of the estimated mean of the ability parameters were as follows: (a) 1.000, (b) 1.000, and (c) 1.000 for 1PL-IRT and 2PL-IRT of the BONE and BRAIN data, indicating almost perfect agreement.

Figures 3–6 show the sampling time for the simulation data of the BONE and BRAIN data. When original-size simulation data were used, the sampling time was shorter in pystan than numpyro CPU version. However, in the simulation data of the BONE and BRAIN data except for the original size, the sampling time was shorter in numpyro CPU version than pystan. Moreover, when the large-sized simulation data (total number of binary responses >30000– 50000) were used, the sampling time was shorter in numpyro GPU version than numpyro CPU version.

**Figure 3.**
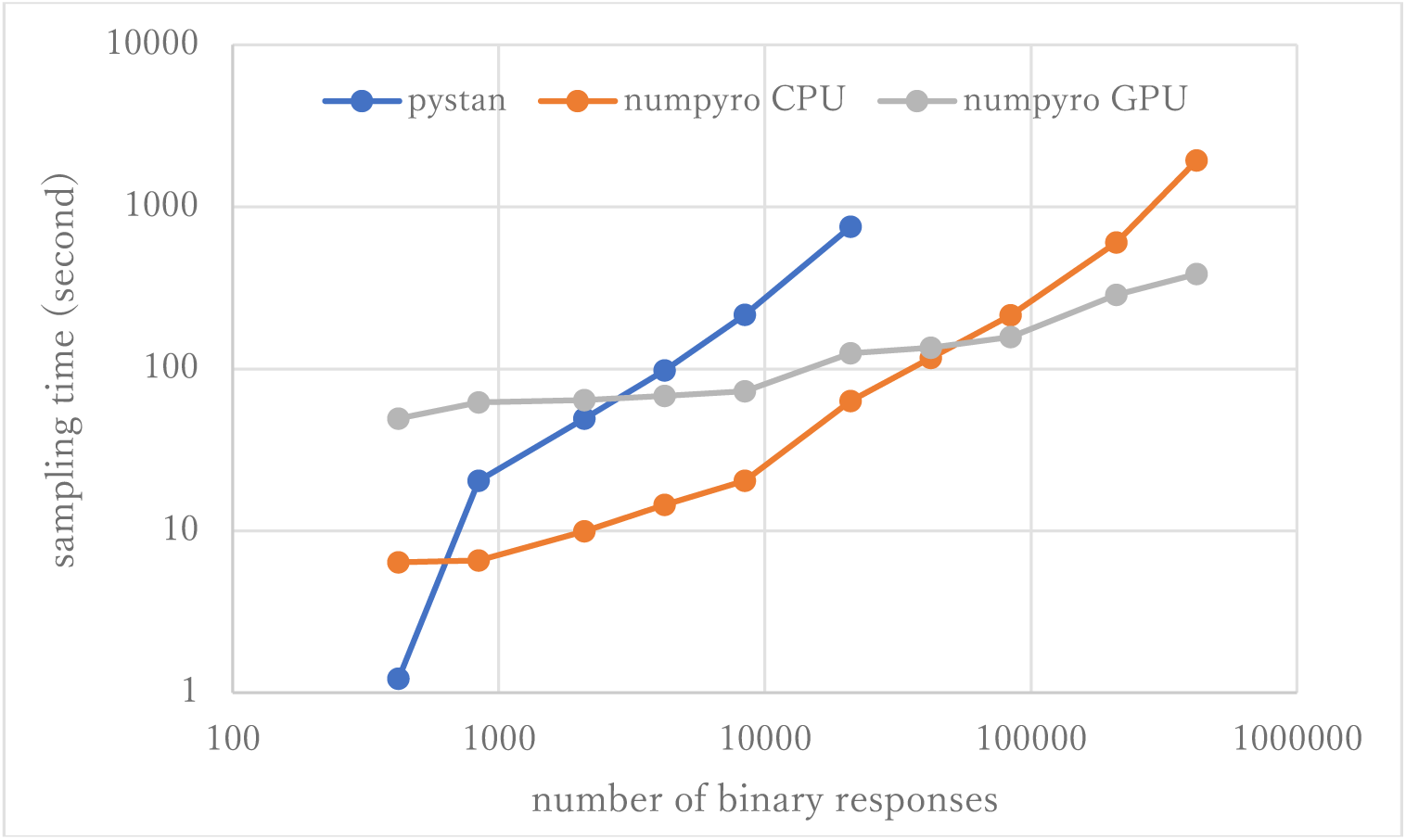
Sampling time of 1PL-IRT for simulation BONE data.

**Figure 4.**
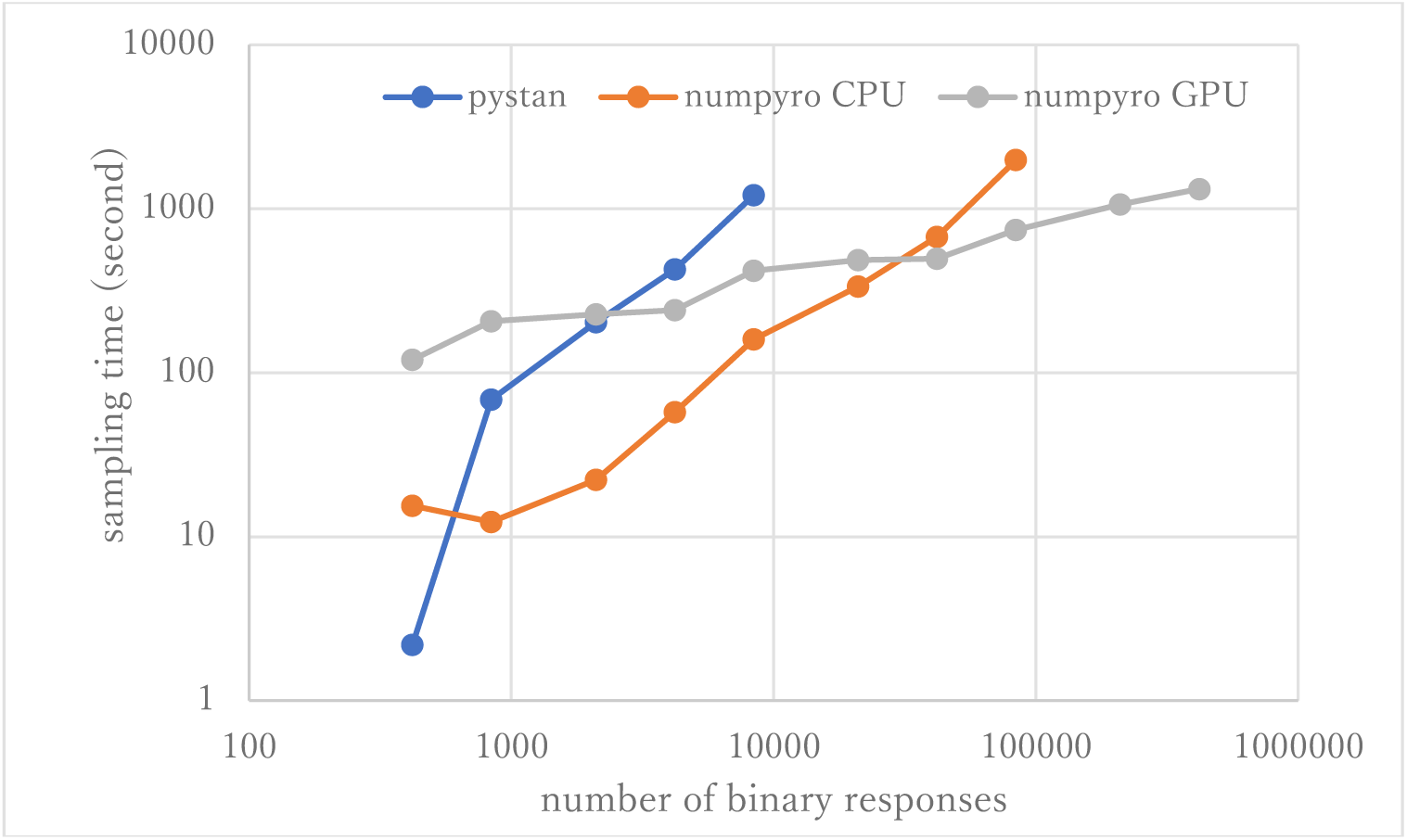
Sampling time of 2PL-IRT for simulation BONE data.

**Figure 5.**
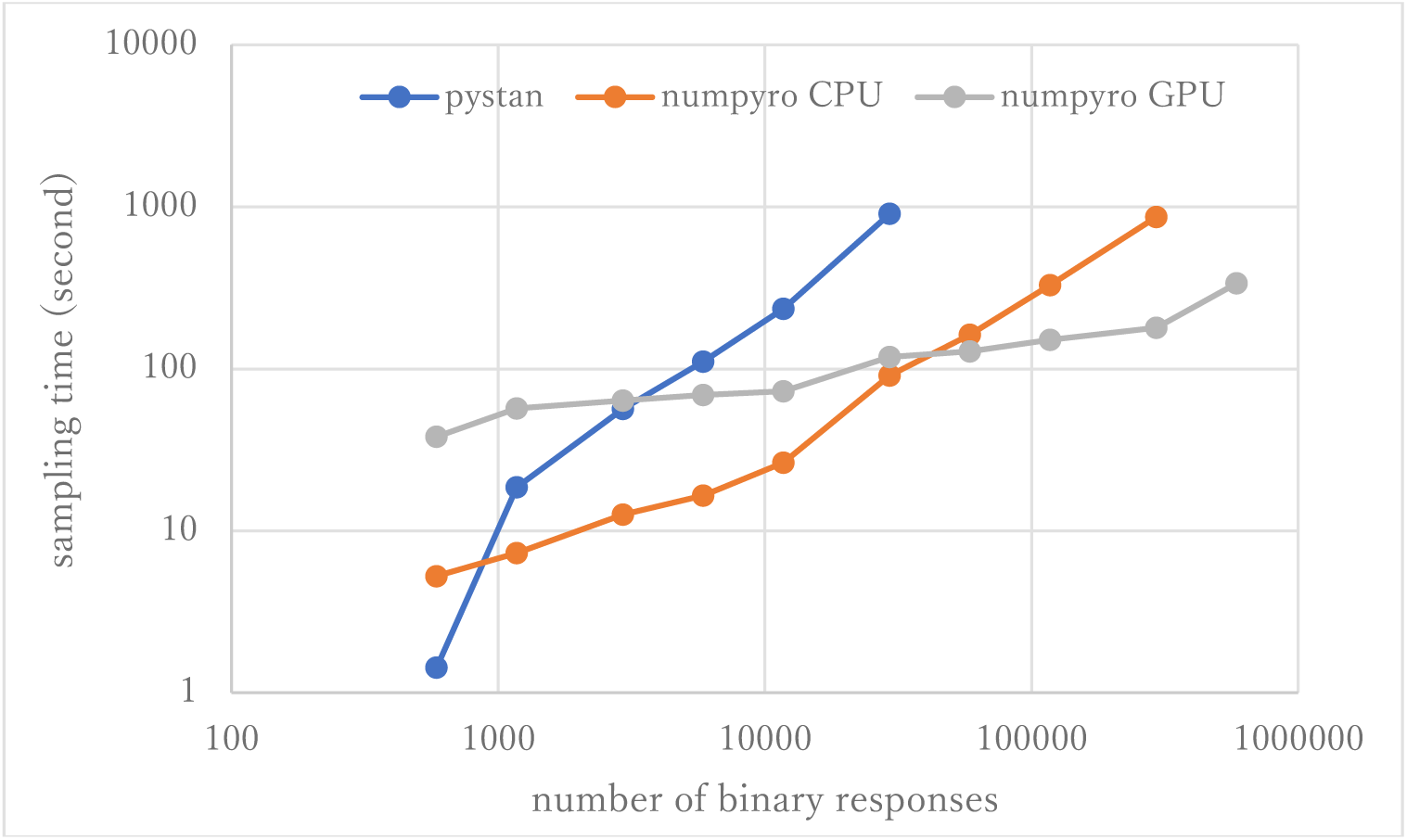
Sampling time of 1PL-IRT for simulation BRAIN data.

**Figure 6.**
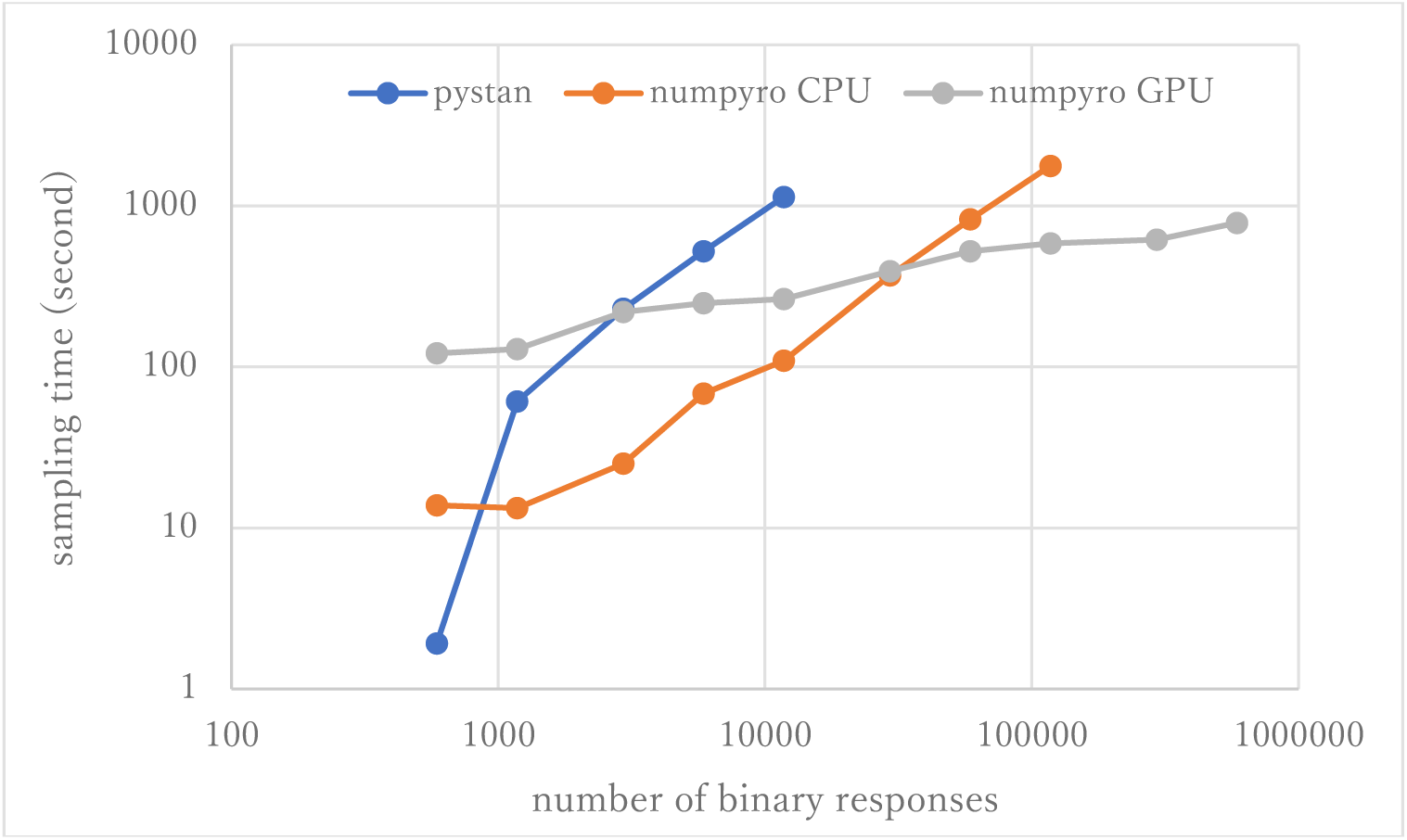
Sampling time of 2PL-IRT for simulation BRAIN data.

## 4 Discussion

The current study aimed to compare the estimation results of the ability parameter of test takers using two different libraries (pystan and numpyro) for two different types of IRT models and medical data (BONE and BRAIN data). The study found that there was good agreement between pystan and numpyro for all the combinations of the IRT models and medical data; there was almost perfect agreement between pystan and numpyro in the CCC values of the estimated mean of ability parameters. The current study also compared the sampling time for the simulation data of the BONE and BRAIN data. We found that while the sampling time was shorter in pystan than numpyro CPU version for the original-size data, it was shorter in numpyro CPU version than pystan for the simulation data except for the original size. For the large-sized simulation data, the sampling time was shorter in numpyro GPU version than numpyro CPU version.

Our results show that there was almost perfect agreement in the ability parameters of the Bayesian IRT between pystan and numpyro. This suggests that researchers can choose either library for implementing the Bayesian IRT. While numpyro requires only Python, both Python and Stan (two different programming languages) are necessary for pystan. Many practitioners and researchers may find numpyro to be simple and straightforward.

Although we used the simulation data, our results of sampling time show that the fastest libraries differed based on the total number of binary responses. Specifically, pystan was the fastest for the original-size simulation data, while numpyro CPU version was the fastest for the small-sized and medium-sized data. For the large-sized simulation data, the sampling time was shorter in numpyro GPU version than numpyro CPU version. This implies that practitioners and researchers should select either pystan or numpyro based on the data size.

Tables 3 and 4 show that the total number of latent parameters may affect the usefulness of GPU for reducing the sampling time. In complex models, such as 2PL-IRT used in this study, numpyro GPU version may tend to be faster than numpyro CPU version. The effect of GPU on the sampling time should be evaluated in future studies.

This study had several limitations. First, we evaluated only Bayesian 1PL-IRT and 2PL-IRT. Further studies are needed to investigate the effectiveness of pystan and numpyro in other types of Bayesian models. Second, although we evaluated the sampling time, we used the simulation data instead of real-world data. Future studies should use real-world data to evaluate the sampling time. Third, we used Google Colaboratory. Although Google Colaboratory has several merits (e.g., ease of use and availability), our experiments were performed using limited types of hardware provided by Google Colaboratory.

In conclusion, the current study demonstrated that both pystan and numpyro were effective in the estimation for 1PL-IRT and 2PL-IRT of the BONE and BRAIN data. Moreover, the study provides useful information about the sampling time for the different data and the libraries using the computer simulations. The results of this study may be helpful for researchers and practitioners who use these models for large-scale data. Overall, the findings of this study contribute to the growing body of research on the application of the Bayesian methods in medical data.

## Data Availability

All data produced in the present study are available upon reasonable request to the authors.

